# Estimation of the Total Number of Infected Cases in the 5^th^ Wave of COVID-19 in Hong Kong

**DOI:** 10.1101/2022.08.12.22278708

**Authors:** W.K. Chow, C.L. Chow, C.H. Cheng

**Affiliations:** Department of Building Environment and Energy Engineering, The Hong Kong Polytechnic University, Hong Kong, China; Department of Architecture and Civil Engineering, City University of Hong Kong, Hong Kong, China

## Abstract

The total number of infected cases in a region in an epidemic is an important measure of the severity of the disease. With the increase in the number of infected people, the number of susceptible people will be reduced, and the recovery number is increased. The present study attempts to estimate the total number of infected cases in the 5^th^ wave of COVID-19 in Hong Kong based on the daily additional cases supplied by the government based on two test schemes. Scheme 1 covers citizens suspected to be infected as referred by medical professionals, or requested or reported by citizens themselves, those returning from overseas, and those in close contact with the infected persons. Scheme 2 covers residents in buildings with a high concentration of virus in sewage. Polymerase chain reaction (PCR) test and then rapid antigen test (RAT) after 26 February 2022 were accepted by the Hong Kong Special Administrative Government in confirming infected cases. The number of infected cases in these two schemes were compared.

A prediction model on infection case was proposed based on the transient daily infection curves. The averaged recovery number was estimated by assuming a 10-day infection period, including an incubation period of 5 days, and another 5 days for recovery. The transient number of infected, susceptible, recovered people were then presented. An adjustment factor to extend the scenario to the whole population of 7 million in Hong Kong was estimated and applied to study infection number in Hong Kong. Further, it appears that the infection number at the later stage of the 5^th^ wave is weak around end July 2022. However, the number stayed at a constant value in comparing with rapid rise at the early stage in February 2022, even though the gathering activities were kept normal.

## 1. Introduction

Rapid outbreak of the 5^th^ wave of COVID-19 in the Hong Kong Special Administrative Region (HKSAR) started from January 2022 [1-3]. This wave of outbreak is a quick incident infecting from 0 to over 1.2 million citizens (over 15% of 7 million) by end May 2022. Appropriate containment schemes have to be operated [4-6]. Catering services and public transport with inadequate ventilation or even with ventilation system turned off [7,8] should be watched. Healthcare systems handling testing, transportation, quarantine accommodation, medical treatment and temporary dead body storage are all overloaded.

The number of infected cases [3,9] is a key parameter to determine actions to take on emergency management such as locking down the city [3,6]. For example, locking down the city by keeping people staying at their unit would cut off or at least delay further infection. The number of infected people can only be determined from appropriate identification tests [10,11].

Restriction-Testing Declaration (RTD) was executed in the HKSAR for controlling the spread of COVID-19 as announced in government press release [12]. Compulsory Testing Notice (CTN) was issued in restricted area including premises like residential buildings. The criteria for making such decisions cannot be reported to the public quickly because infection changes fast. There are a number of transient factors, depending on the severity of the pandemic or epidemic, the estimated number of contracted cases in the specified area, the testing capacity, and the manpower available as stated in [13]. Examples are those suspected to be infected by themselves or medical doctors; those returning from overseas; and buildings with confirmed cases detected in at least two units within a short period of time are included.

In addition to monitoring the number of confirmed cases in a building, the government also makes decision based on another scheme related to sewage surveillance. Regular tests on sewage samples were carried out in collaboration with a university for identifying coronavirus in all districts. RTD with CTN issued or simply CTN issued were based [14] on the local test results in conjunction with other factors.

Laboratory polymerase chain reaction (PCR) tests are used. Rapid antigen tests (RAT) are accepted after 26 February 2022 because there are too many people to be tested every day. Further, the government had purchased adequate RAT tests. But there are doubts on the results and RAT results must be confirmed with PCR tests. Correlation between the positive rates of the above two batches of tests will be studied in this paper. Estimating daily infection from the two sets of tests can give a hint on how many citizens are infected every day.

There are different estimations on the number of infected cases in the whole city. Some predicted value is over 4 million. If the infected cases increased from 1000 to 1 million takes a few weeks, then 4 million of infected citizens infect the other citizens not yet infected (7 – 4 = 3 million citizens) would happen quickly.

All these points will be discussed in this paper. Finally, the changes of infection number during the beginning in February 2022 and at the later stage in July 2022 of the 5^th^ wave are compared.

## 2. Daily Tests

As discussed in above, the criteria of exercising CTN or RTD based on sewage testing results or number of confirmed cases cannot be announced quickly to the public. This is because the daily situation changes rapidly, leading to having many factors varying with time. Even the threshold viral load that determines the compulsory testing order CTN or RTD have to be changed frequently, depending on the outbreaks. No other places can respond so quick and announce so fast such as in Hong Kong. Note that the threshold would be changing rapidly with the extent of infection. The number of buildings tested under CTN per day due to previously reported positive cases [15] are 10 buildings per day from 22 to 24 March 2022; to 25 buildings per day from 31 March to 2 April 2022. Around 40,000 people were tested every day.

Further, sewage surveillance would provide data on whether residents in the building tested are infected by COVID. Routine testing on sewage in buildings or buildings suspected to have asymptomatic carriers would give some useful data on the infected number. An example case study of sewage surveillance on Delta variant outbreak in Hong Kong was reported by Deng et al. [16]. Routine testing of a sewage sample on 21 June 2021 gave a high viral load of SARS-CoV-2. All residents in that building were tested on 23 June 2021 with one case confirmed on 24 June 2021. After that, zero cases were found for 7 months.

CTN are required when the criterion of having more than some (over 2) cases is met. However, CTN are also required when the virus concentration in the sewage of the building(s) reaches a threshold. In summary, two daily test schemes [2,17,18] are practicing for identification of infection:

- Scheme 1: This is the main scheme with CTN requesting citizens having close contacts with infected persons, people returning from other areas and those suspected to be infected with similar symptoms to carry out compulsory PCR identification tests. Over 100,000 PCR tests were done every day. After 26 February 2022, rapid antigen test (RAT) results [17] were accepted with positive results W_1_ (PCR only) and W_2_ (Accepted RAT after 26 February 2022) as shown together in Fig. 1a.

**Fig. 1:**
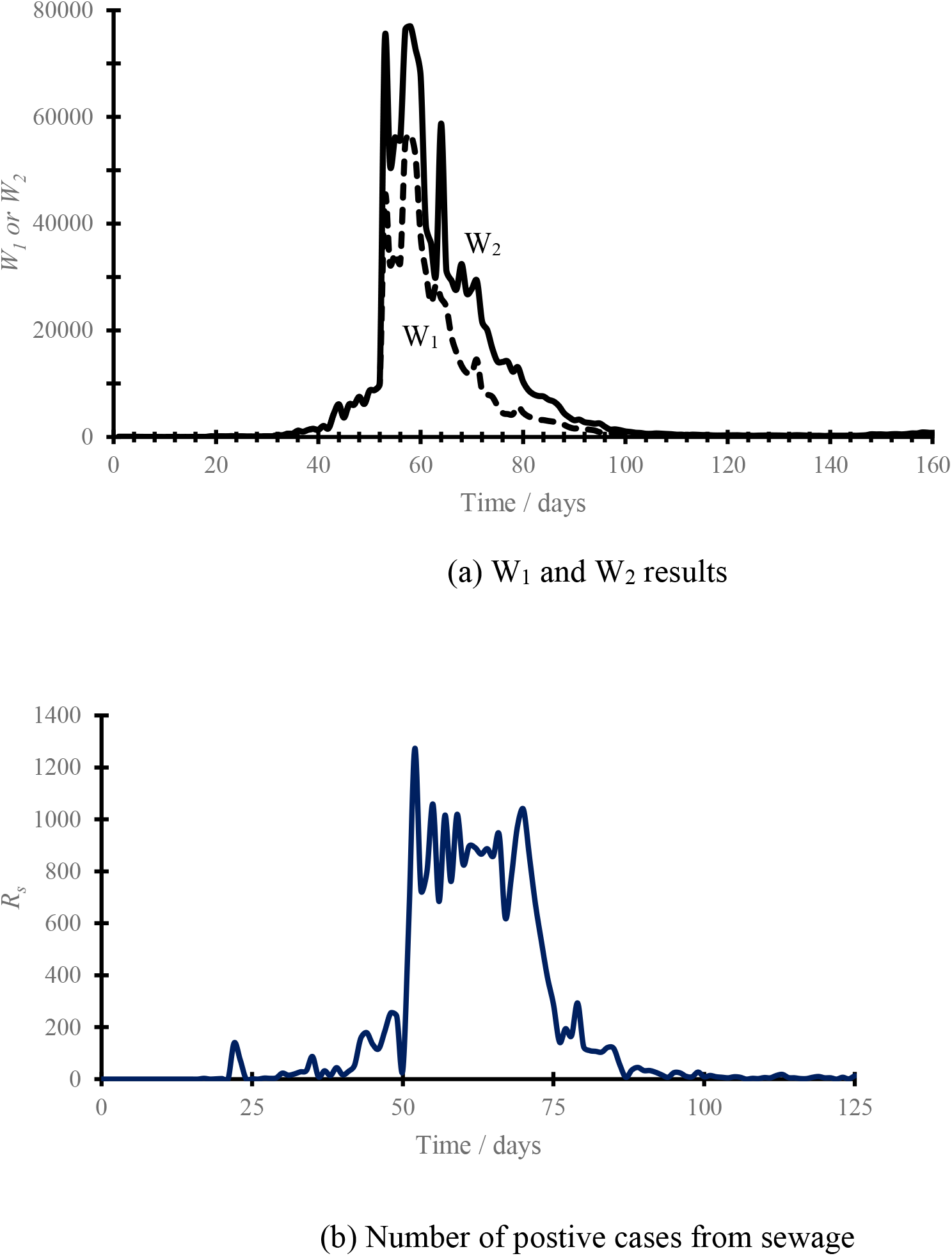
Daily test results. Reference: Government of the Hong Kong Special Administrative Region. http://www.gov.hk/
- Scheme 2: Residents in buildings have sewage contaminated with virus are required to do PCR tests. The results [17] on positive number R_s_ for compulsory PCR testing under RDT with contaminated sewage are plotted in Fig. 1b. Buildings tested as in the table must have sewage contaminated. Otherwise, CTN will not be issued to residents of the buildings.

The number of buildings with sewage contamination tested a day was not announced. However, there must be some criteria set up by the Health Department but not reported nor discussed before at their daily TV media interviews. The number of cases recorded in buildings over the past 7 days might be exceeding 50, as searched from news, with some from government website [18].

## 3. Testing Results

Taking the daily number of positive cases R_s_ for residents in buildings with sewage contaminated as in Scheme 2, the total number of PCR tests under CTN in Scheme 1 is W_1_, and the total number of PCR plus RAT reported by citizens after 26 February 2022 is W_2_. All these daily recorded values of confirmed positive cases, W_1_, W_2_ and R_s_ are plotted in Fig. 1a and b.

Two linear lines are fitted for W_1_ with correlation coefficient of 0.9055 and W_2_ with correlation coefficient of 0.8459:

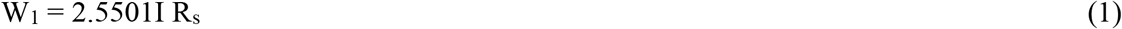

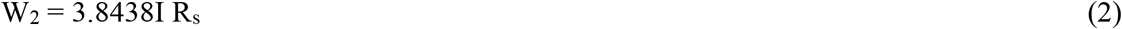

These two linear lines for W_1_ and W_2_ with R_s_ are plotted in Fig. 3a.

**Fig. 2:**
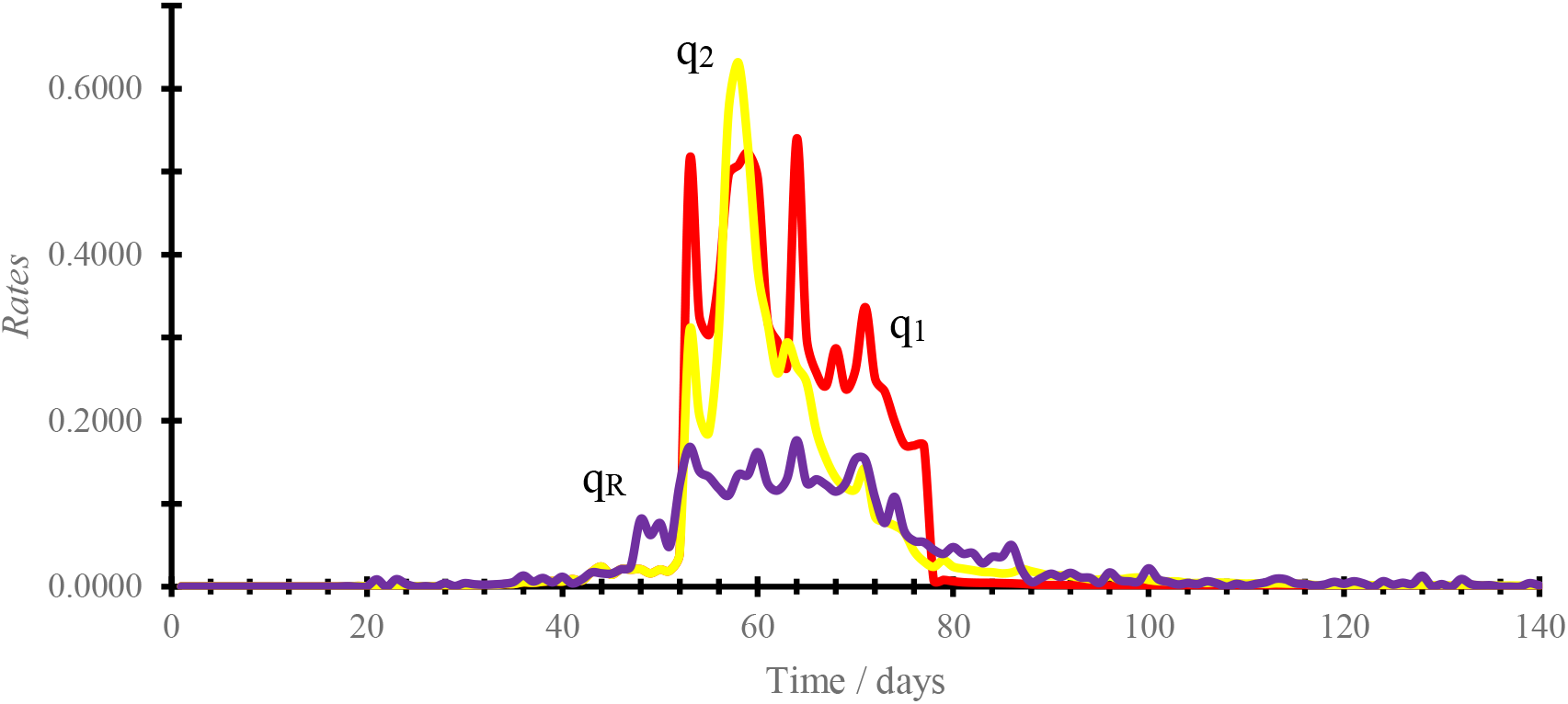
Comparing rate of 3 tests. Reference: Government of the Hong Kong Special Administrative Region. http://www.gov.hk/

**Fig. 3:**
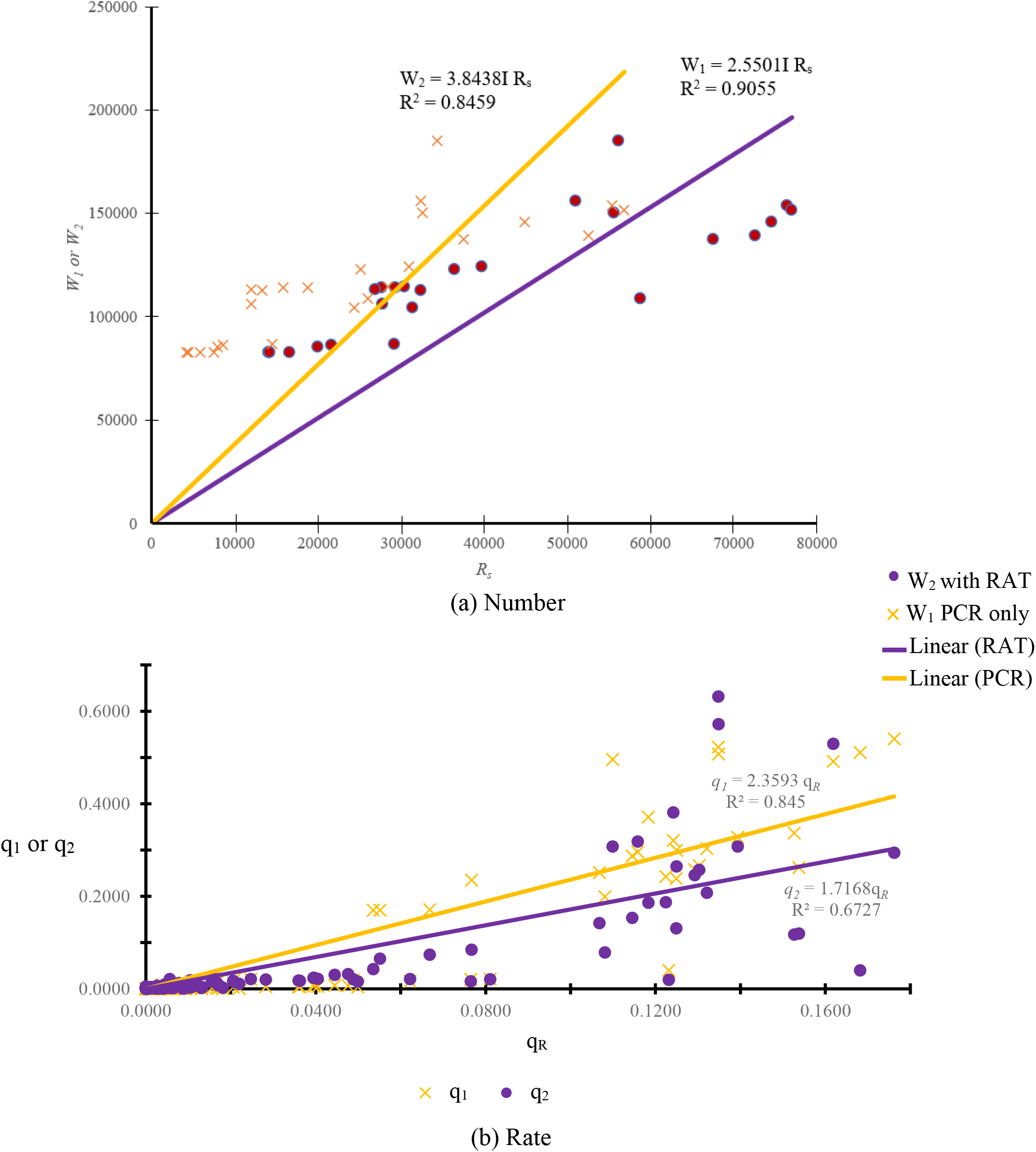
Correlation between the two tests.

From these three sets of data on the number of positive cases W_1_, W_2_ and R_s_, the rate q_1_, q_2_ and q_R_ are computed from the total number of PCR tests T_1_ under Scheme 1, and total number of RTD tests T_R_ as follows:

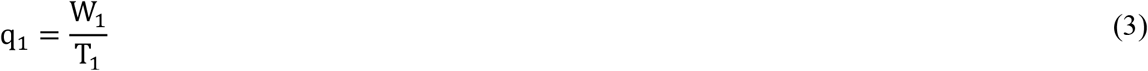

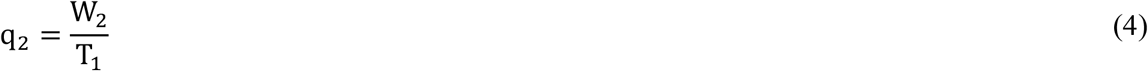

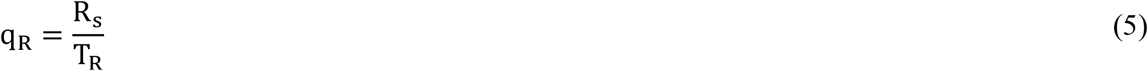

Results of q_1_, q_2_ and q_R_ are plotted in Fig. 2. Results of q_1_ and q_2_ plotted against q_R_ are shown in Fig. 3b.

The one for q_1_ with q_R_ has a correlation coefficient of 0.845 and the one for q_2_ with a lower correlation coefficient of 0.6727:

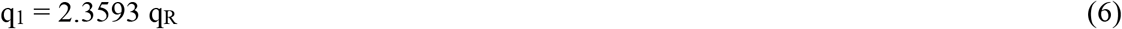

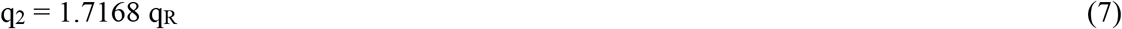

The following points are observed from the figure:

- Rate of positive tests q_R_ from sewage contaminated buildings is related to the rate for total PCR tests q_1_ with a fairly high corelation coefficient. It is not too clear with q_2_ with a low correlation coefficient. A possible reason is that RAT tests are not so accurate. Further, these tests were carried out by residents themselves only, not by trained technicians.
- As q_1_ is related linearly to q_R_, the value of q_R_ can be used to estimate q_1_ if the number of buildings testing sewage N_s_ is known.
- The total number of PCR tests done every day is also important to get the percentage of infection.

Testing residents with PCR in buildings with sewage contaminated is good. However, the daily number of buildings N_s_ with sewage tested must be known to get the denominator as the total number of residents involved N_B_ for estimating rate R_s_/N_B_ (not just R_s_ divided by the number of residents involved in contaminated buildings, t_R_) of infected cases in the tested buildings.

Sewage contaminated buildings can then give an estimate on daily total number of infected cases in the whole city with appropriate adjustment.

## 4. Estimation of Different Numbers

S(n), I(n), R(n) are the numbers of the susceptible, infectious, and recovered people in modelling [20-22] and instantaneous numbers at time of the n^th^ day. They are related to the total number of people N_o_ by:

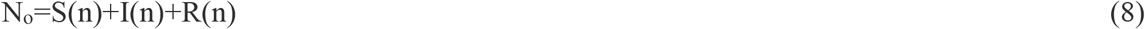

The above equation has to be applied carefully with the symbols used in a proper sense. That is because the model holds for large number of S, I and R. Predictions on S, I and R have larger deviations at the initial stage for this kind of stochastic models [6]. Recovery comes from infection first as a series of events. Some cases of recovery and infection appear to happen in parallel because infection happens at different time, and the duration of infection might be different for different people. These two reasons lead to infection and recovery process happen at the same time, though the numbers are different.

Note that the total number of R(n) is lower than or equal to the total number of I(n), or

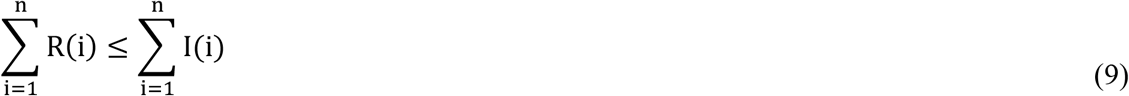

However, only the daily number of additional confirmed cases I(n) is reported every day. The total cumulative number of infection CI(n) is:

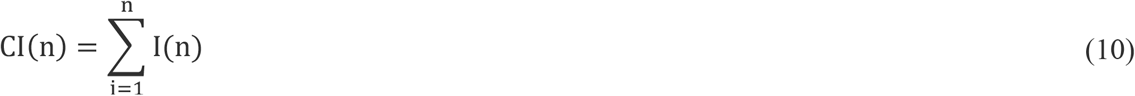

However, it takes some time for people to recover if they can survive. It is difficult to have a fixed recovery day. Taking 10 days to be the maximum recovery period for infected person is reasonable. Recovery number R(n) on the n^th^ day is:

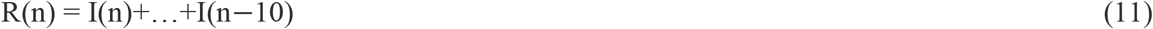

The total number of citizens still have infected symptoms IT(n) on the n^th^ day after deducting recovery:

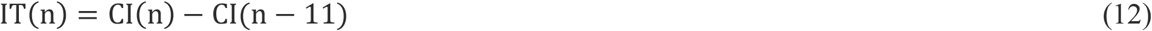

As IT(n) and R(n) are known, S(n) can be calculated from equation (8).

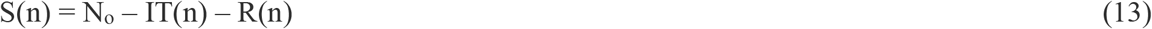

The curves S(n), IT(n) and R(n) deduced from I(n), and N_0_ (7 million) are plotted together with I(n) in Fig. 4.

**Fig. 4:**
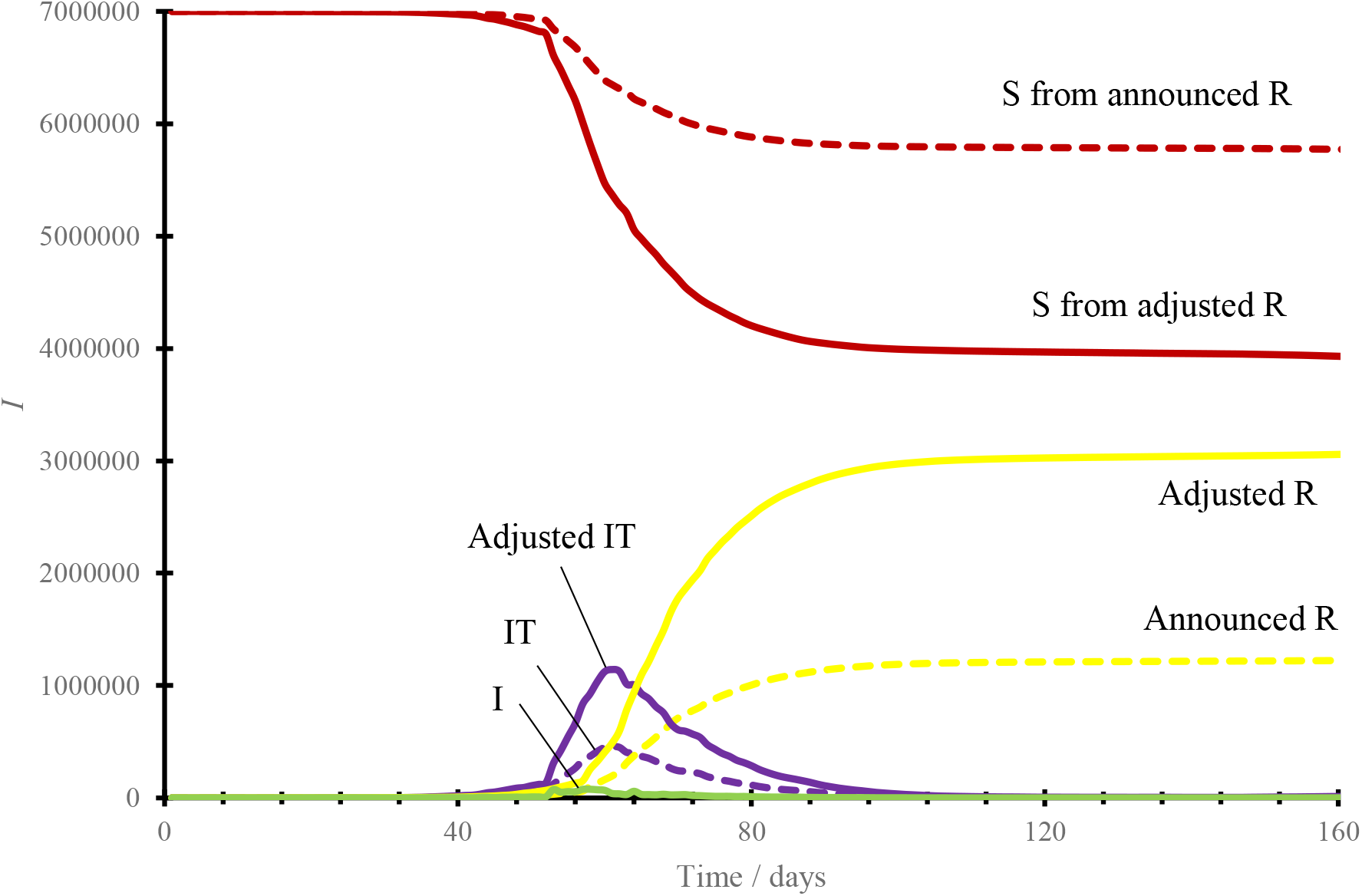
SIR curves.

## 5. Adjustment Factor

There are different views on estimating the infection number I. The Chief Executive (CE) held daily updating meetings during the 5^th^ wave of outbreak when the daily infection number was over 1000 since March 2022. At the CE media meeting on 8 March, 2022 [19], the infection rate picked up in a specified premise of compulsory testing with contaminated sewage was 4.9% (or 5%). Adding the earlier confirmed cases of 20% would give a total of 25% infection. Using this figure to extrapolate the approximated number of Hong Kong infected cases, 7 million times 25%, giving a minimum estimation of 1.75 million infection. If each person infected another 2 family members, this matches with another predicted infection cases of over 5.25 million from the 1.75 million confirmed cases.

However, not many buildings have sewage contaminated under Scheme 2 above. Those buildings taken out to have compulsory test might be only 1% (a guessed value only, government should have kept data) of the total number of buildings in Hong Kong. The instantaneous number of people infected is only 25% maximum in a sewage contaminated building, multiplying 1.8 million by 1% gives 18,000 people infected. However, a cycle of infection might take a maximum of 10 days, so the new infection is only at most 180,000 in a cycle of 10 days.

To estimate the total number of daily infected cases on Scheme 1 with PCR only, the total number of tests might be around 0.5 million maximum, where T_1_ is much less than the total number of citizens of 7 million. The total number of PCR tests varied every day, but still a large number of people are either not infected or without any symptoms.

Taking a rate 0.4 as in Fig. 2, multiplying the total number of infected IT(n) on the n^th^ day by 1/0.4 or 2.5 gives an estimated total number of infections.

All values of S and R are adjusted as in Fig. 4. This is only one of the possibilities.

## 6. At the Later Stage

The number I increased at the later stage of the 5^th^ wave [17] from 1 July to 1 August 2022. The daily number was kept at around 4,000 and compared with the beginning as shown in Fig. 5. There are differences in the increase of I at the beginning of the 5^th^ wave.

**Fig. 5:**
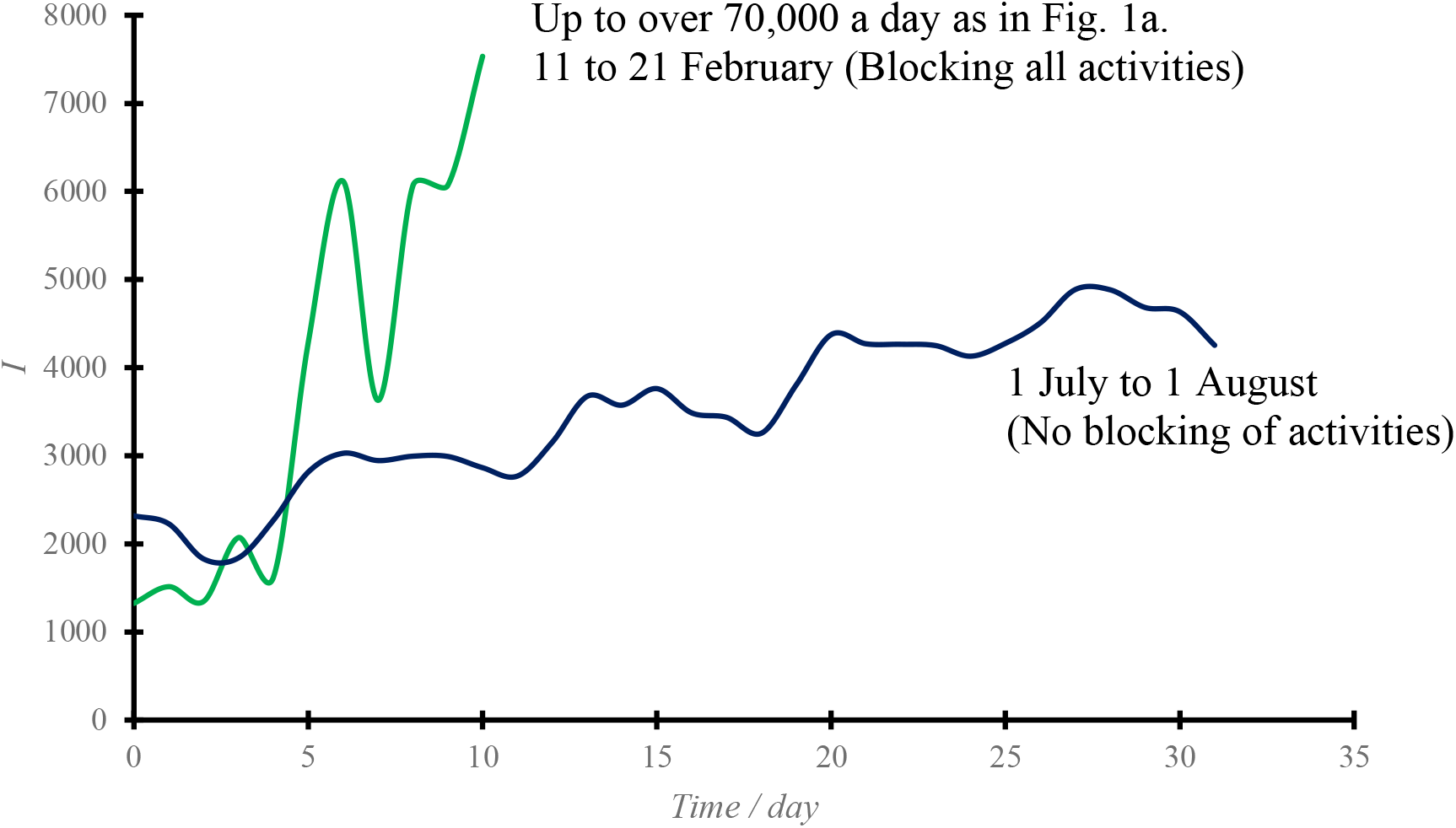
Comparison of beginning and end of 5^th^ wave. Reference: Government of the Hong Kong Special Administrative Region. http://www.gov.hk/

Two points are observed from the figures:

- At the beginning around end February 2022, there was tight control on gathering activities such as imposing dinning restrictions in restaurants, ordering pubs to be closed and suspending many other functions. Face-to-face school classes were also suspended and moved to online classes. Even so, the values of I increased very fast in the beginning from 11 February to 21 February 2022 as in Fig. 5.
- However, almost all gathering activities resumed normal with close human contact at end July. Further, there are fewer number of people having serious symptoms. The healthcare system is still functioning properly up to 9 August 2022.

A possible reason is that the residents might have built a herd immunity barrier for protection against COVID-19. Further investigation is suggested.

## 7. Discussions

The daily number of infected cases in a city should be estimated properly for the government to make appropriate decision on emergency management. Instruction to lock down the city [3,6] is an example, which is difficult to decide. Early lockdown is effective in achieving objectives on mitigation and suppression of outbreaks. Containment is necessary [5,23] with quick identification tests [10,11,24-26]. Air transmission should be watched [7,27].

However, economic impacts might lead to criticisms, some businesses with other cities with difficulties to arrange lockdown are forced to be terminated. Air traffic, catering services and tourist industry suffered most. Further, the medical system on handling emergency can be better allocated. As experienced in March 2022, the elderly suffered. The government must compromise whether those businesses with gathering activities should be opened.

The number I can be obtained through regular mass screening of the people in the region. However, mass screening involves immense resources and seriously affects people’s daily life and jeopardizes the local economy, leading to negative societal image and huge economic loss. In view of this, estimation rather than direct acquisition of the total number of infected cases has been proposed, based on the information available.

## 8. Conclusions

The following can be observed from the above study:

- The daily infected number recorded from the two schemes of identification tests on confirmed cases are linearly correlated. The correlation is better for taking PCR tests only.
- Knowing the positive testing rate of sewage can give an estimation of the positive rates of PCR tests.
- The total number of infected persons on a certain day can be estimated from the observed recovering period for those infected, say 10 days maximum.
- The recovery number can be deduced from the total infected number on a certain day.
- The total number of recovery, and susceptible in the whole city can be adjusted from the observed rate of identification tests.
- The daily infected number change rate at the beginning and at the later stage of the 5^th^ wave due to a COVID-19 variant is different.
- It is observed that at the beginning, the daily infection number I increased from 4,000 to over 70,000 quickly as in Fig 1a. At the later stage of the 5^th^ wave, the number I was kept at about 4,000 and increased slowly. Even taking 10 days on recovery, the total number of infections on a certain day in August is around 40,000, much less than those number around March.

## Data Availability

All data produced in the present work are contained in the manuscript

## Notes

### Competing Interest Statement

The authors have declared no competing interest.

### Funding Statement

This study did not receive any funding

